# Analgesic Efficacy and Safety of Caudal Epidural, Dorsal Penile Nerve, and Pudendal Nerve Blocks in Hypospadias Repair Surgery in Children: Updated Meta-Analyses

**DOI:** 10.1101/2025.07.19.25331825

**Authors:** Moumen Arnoaut, Omar Chikh Amine, Mohamad Jaber Arab, Abdullah Aladnan Aljammas, Mohamad Amin Kreid, Abdulmajid Makki, Kais Alissa, Mohammed Al-mahdi Al-kurdi, Mohammad Tammam Saffour, Mohamad Morjan

**Author notes:** Corresponding Author. Moumen Arnaout Faculty of Medicine, Aleppo University, Aleppo, Syria.

## Abstract

**Background:** Hypospadias affects 1 in 150-300 male births and requires surgical correction. Optimal regional anesthesia technique selection is crucial for pain management in these pediatric procedures.

**Objective:** To compare the analgesic efficacy and safety of caudal epidural (CB), dorsal penile nerve (DPNB), and pudendal nerve blocks (PNB) in children undergoing hypospadias repair surgery.

**Evidence Review:** We conducted a systematic review by searching through PubMed, CENTRAL, SCOPUS, Web of Science, EBSCOhost, and clinical trial registries for randomized controlled trials on 26 November 2024. Studies comparing dorsal penile nerve (DPNB), or pudendal nerve blocks (PNB) and caudal epidural Block (CB) in children with hypospadias were included. RevMan v5.4. was used to conduct two pairwise meta-analyses, pooling dichotomous data using risk ratio (RR) and continuous data using mean difference (MD) with a 95% confidence interval (CI). Quality was assessed using the Cochrane Risk of Bias 2 tool.

**Findings:** Ten randomized trials (768 patients: CB=337, DPNB=173, PNB=169) were included. Six studies had some concerns for bias, no studies were identified as carrying a high risk of bias. PNB demonstrated significantly lower pain scores compared to CB at 6 hours (SMD: -0.50 with 95% CI [-0.77, -0.23], P=0.0003), 12 hours (SMD: -1.95 with 95% CI [-3.29, -0.61], P=0.004), and 24 hours (SMD: -1.20 with 95% CI [-2.29, -0.11], P=0.03) with reduced analgesic consumption (RR: 0.23 with 95% CI [0.05, 0.99], P= 0.05). No significant differences in pain scores were found between DPNB and CB. DPNB was associated with decreased hypotension compared to CB (RR: 0.27 [95% CI: 0.08, 0.93], P=0.04).

**Conclusions:** For pediatric hypospadias repair, PNB provides superior postoperative analgesia compared to CB, while DPNB offers comparable pain control with improved hemodynamic stability. Regional block selection should balance analgesic efficacy with safety profile. More clinical trials that provide homogenous and no biased data is needed.

## 1. Introduction

Hypospadias, a congenital abnormality affecting male external genitalia, is characterized by the atypical development of the urethral fold and ventral foreskin of the penis [1]. It is a relatively common condition, with incidence rates ranging from one in every 150 to 300 live births [2]. Recognizing the importance of adequate pain management for pediatric surgical procedures, the World Health Organization recommends analgesia for all children undergoing such operations [3].

The innervation of the penis involves the pudendal nerve, which originates from the sacral plexus in the pelvis, with several branches, including the dorsal nerve of the penis [4]. Among the anesthesia methods utilized in surgeries for hypospadias repair are Dorsal Penile, Pudendal, and Caudal Epidural blocks, each with its specific technique and benefits [5-6].

Caudal epidural block (CB) is frequently employed in children for surgical anesthesia, involving the insertion of a needle through the sacral hiatus to access the sacral epidural space [7]. On the other hand, the Dorsal Penile Nerve Block (DPNB) is commonly used to provide regional anesthesia for male circumcision, effectively numbing the penile area with minimal local anesthetic volume [8-9]. Pudendal nerve block (PNB) is another anesthesia technique, widely utilized for postoperative pain relief in conditions like hemorrhoids, achieved through the injection of a local anesthetic around the pudendal nerve trunk [10-11].

From the above, it is evident that hypospadias is a common congenital condition among newborn males, leading to potential future deformities that may impede their quality of life. Consequently, corrective surgical intervention is essential to prevent such complications [1]. However, the surgical procedure itself carries inherent risks, some of which may be attributed to the type of anesthesia. Despite the availability of these anesthesia methods, there is currently no definitive evidence favoring one approach over the others. The objective of this systematic review and the two pairwise meta-analyses is to compare the efficacy and safety of dorsal penile nerve, pudendal nerve, and caudal epidural blocks in hypospadias repair surgeries in children. By evaluating the resulting complications and the effectiveness of postoperative pain management, our study aims to provide insights into selecting the most suitable anesthesia method for infants undergoing such procedures.

## 2. Material and Methods

### 2.1. Protocol Registration

The study’s protocol was registered in PROSPERO under ID CRD420251057876 following the Preferred Reporting Items for Systematic Review and Meta-analysis of Interventional Studies (PRISMA) statement [12], and the Cochrane Handbook for Systematic Reviews and Meta-Analysis [13] guidelines.

### 2.2. Data Sources & Search Strategy

Databases, including PubMed, CENTRAL, SCOPUS, Web of Science, EBSCOhost, Clinicaltrial.gov, and International Clinical Trials Registry Platform (ICREP), were searched through 26 November 2024 using a search strategy of ((Pudendal OR Penile OR DPNB OR "Dorsal Nerve" OR Caudal OR Epidural) AND (Hypospadias OR Hypospadia)). No restrictions on publication date or geographical area were made. More details are in **(Table S1).**

### 2.3. Eligibility Criteria

Randomized controlled trials that met our PICO criteria were included: population (P): male children (≤ 18 years) undergoing surgical repair of hypospadias; intervention (I): Peripheral blocks including pudendal nerve block or dorsal penile nerve block; comparison (C): caudal epidural block; outcomes (O): our primary outcome was postoperative pain score. Our secondary outcomes in the PNB vs. CB. comparison include: analgesic consumption within 24 hours postoperatively, block procedure time, surgery time, and total dose of acetaminophen, while our secondary outcomes in the DPNB vs. CB. comparison include: surgery time, and safety outcomes including: hypotension, bradycardia, and vomiting. Observational studies, duplicates, reviews, pilot studies, case reports, editorials, letters, commentaries, conference abstracts, non-English language studies were excluded.

### 2.4. Study Selection

Six reviewers independently screened the titles and abstracts using the Rayyan platform. After excluding duplicates, reviewers independently screened the full texts in accordance with the previously mentioned eligibility criteria. Any conflicts have been resolved by consensus.

### 2.5. Data Extraction

Four reviewers independently extracted data from the eligible studies using an Excel sheet. This Was divided into three main sections: (1) a summary sheet (Country, time frame, total population, intervention and comparison including the type of block and medication used, pain assessment scale, outcomes, and study conclusion); (2) baseline characteristics (number of patients in each group, age, weight, and height); and (3) study outcomes (surgery time, block procedure time, pain scale, total dose of acetaminophen, analgesic consumption within 24 hours postoperative, hypotension, vomiting, and bradycardia). Conflicts were discussed and resolved by consensus.

### 2.6. Risk of Bias

The risk of bias assessment was implemented by two independent reviewers according to Cochrane’s risk of bias assessment tool 2 and considering the following domains: Random sequence generation (selection bias); Allocation concealment (selection bias); Blinding of participants (performance bias); Blinding of outcome assessor (detection bias); Incomplete outcome data (attrition bias); Selective reporting (reporting bias). Reporting bias was assessed by screening clinical trial registers through the comparison of outcomes defined in the protocol with each published study. Conflicts were resolved by consensus.

The risk of bias in each study was defined as below: Low risk (low for all main domains), unclear risk (unclear for one or more main domains), and high risk (high for one or more main domains).

### 2.7. Statistical Analysis

We performed two pairwise meta-analyses using RevMan v5.4. Continuous outcomes were pooled as mean difference (MD) between the study groups from baseline to endpoint, while categorical/dichotomous outcomes were pooled as event/total in each study group and represented as risk ratio (RR), both with a 95% confidence interval (CI). The random-effects model was used only where there was considerable heterogeneity. Chi-square and I-square tests were used to evaluate heterogeneity, where the Chi-square test detects the presence of heterogeneity, and the I-square test evaluates its degree. I-square was interpreted according to the Cochrane Handbook [13] as follows: heterogeneity is not significant for 0–40 percent, moderate for 30–60 percent, substantial for 50–90 percent, and considerable for 75–100 percent. Heterogeneity was considered significant when an alpha level was below 0.1 for the Chi-square test. To address heterogeneity, a leave-one-out sensitivity analysis was performed by iteratively excluding each study one time from the pooled analyzed studies.

## 3. Results

### 3.1. Search Results and Study Selection

After searching the following databases (PubMed, CENTRAL, SCOPUS, Web of Science, EBSCOhost, Clinicaltrial.gov, and International Clinical Trials Registry Platform (ICREP)), our search strategy resulted in 9459 records. After duplicate removal, we reached 5904 for title and abstract screening and 351 records eligible for full-text screening. Finally, we included 10 studies by our eligibility criteria **(Figure 1).**

**Figure 1:**
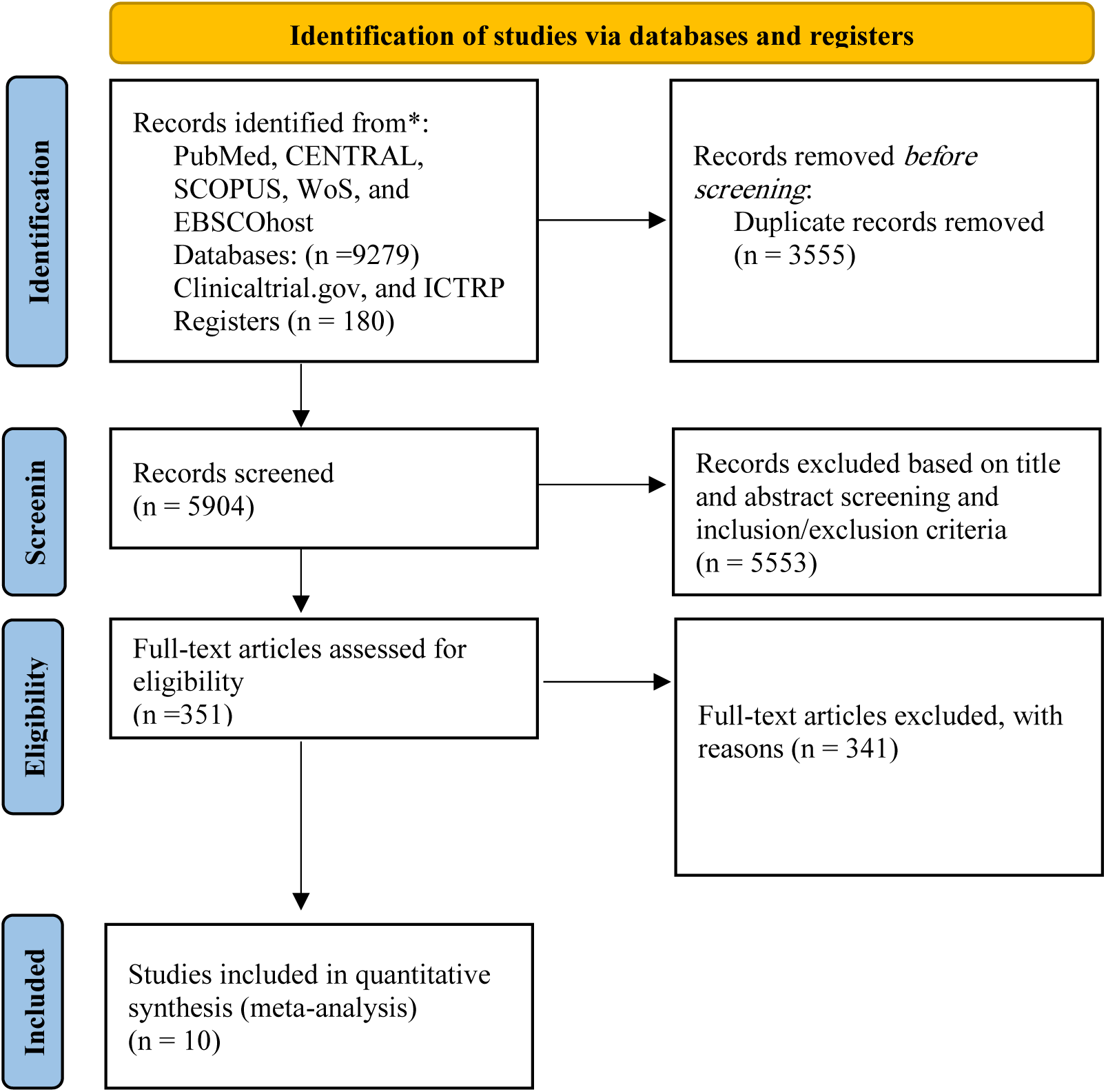
Flow diagram of the study selection process. Wos, Web of science; ICTRP, International clinical trials registry platform.

### 3.2. Characteristics of Included Studies

We included 10 RCT studies [14-23] with 768 patients, with 169 patients in the PNB group, 173 in the DPNB group, and 337 in the CB group, conducted across eight countries. The majority of the included studies assessed the primary outcome, the analgesic efficacy in pain control through various pain scales. More details about the characteristics of included studies and enrolled patients are summarized in **(Tables 1, 2).**

**Table 1.** Summary data of the included studies. ASA: Physical status classification of the American society of anesthesiology; C: Concentration; CHEOPS: Children’s Hospital of Eastern Ontario Pain Scale; FLACC: The face, legs, activity, cry, and consolability scale; FPS-R: Faces pain scale-revised; PACU: Post-anesthesia care unit; OPS: objective pain/discomfort scale; PNB: Pudendal nerve block; CB: Caudal epidural block; DPNB: Dorsal penile nerve block; VAS score: Visual analogue scale.

| Study | Country | Time Frame | Total Population | ASA | Intervention |  | Control |  | Pain scale | Outcomes | Study Conclusions |
| --- | --- | --- | --- | --- | --- | --- | --- | --- | --- | --- | --- |
|  |  |  |  |  | Type of Block | Medication (C) | Type of Block | Medication (C) |  |  |  |
| Ahmed et al 2024 | Pakistan | March to September 2022 | 68 | I-II | Ultrasound-Guided pudendal block | 0.25% bupivacaine 0.2ml/kg | Caudal block | 0.2mg/kg of bupivacaine 0.25% | CHEOPS | Time of surgery, Pain assessment, Complications, and Requirement for Analgesia | Ultrasound-guided PNB is superior to CB in providing adequate perioperative analgesia with fewer complications and decreased requirement of additional analgesic supplements. |
| Ahmed et al 2023 | Sudan | April 2017 to September 2017 | 60 | I-II | Penile block | 0.25% bupivacaine 0.1 ml/kg | Caudal block | 0.25% bupivacaine 0.5 ml/kg | FLACC | Vomiting | The penile block seems to be more effective in decreasing post-op pain, delaying the onset of pain, reducing the need of analgesic rescue, and has milder side effects |
| Elbadry et al 2023 | Egypt | April 2022 to August 2022 | 120 | I-II | Penile block | Bupivacaine 0.5% 0.1ml/kg | Caudal block | Bupivacaine 0.25% 1 ml/kg | FLACC pain scale | Analgesic duration, Pain score, Analgesic requirement, and Incidence of complications | CB is associated with better pain control compared to the penile block / the hypotension incidence was significantly higher in caudal than other techniques. |
| Khalil et al 2022 | Egypt | NA | 150 | NA | Penile block | Total of 4 ml of 0.25% bupivacaine | Caudal block | 0.5 ml/kg of 0.25% bupivacaine | FPS-R | Postoperative pain assessment, Frequency of any intraoperative or postoperative problems. | DPNB may be used to achieve the same analgesic effectiveness as the CB, in pediatric cases having hypospadias corrective operation. Although the CB group had better hemodynamic stability with early recovery from anesthesia. |
| Choudhry et al 2022 | USA | May 2017 to December 2019 | 57 | I-II | Nerve stimulation guided pudendal | Ropivacaine 0.2% 0.25 ml/kg | Caudal block | Ropivacaine 0.2% 1 ml per kg | FLACC pain scale and parental | Postoperative pain score and Block duration | Comparable pain relief is obtained intraoperatively with block alone and postoperatively with a multimodal approach involving NSAID in conjunction |
|  |  |  |  |  | al block |  |  |  | assessment scale |  | with both PNB and CB for analgesia in the first 24h after surgery |
| Karami et al 2021 | Iran | July 2019 to March 2020 | 50 | I-II | Penile block | 0.25% bupivacaine 0.1 ml/kg | Caudal block | One milliliter 0.2% bupivacaine | FLACC | Pain assessment | According to results obtained from this study, hypospadias repair in pediatrics using CB can provide longer analgesia for the patient |
| Ahmed et al 2021 | Egypt | NA | 50 | I-II | Ultrasound-guided pudendal nerve block | 0.3 ml/kg 0.25% bupivacaine | Caudal block | 1 ml/kg 0.25% bupivacaine | The "objective pain scale" | Postoperative pain, Analgesic requirement, Time needed to perform the block, and Duration of surgery | The current study revealed that US-guided PNB provided significantly prolonged postoperative analgesia, reduced postoperative analgesic requirements as compared with CB in pediatric patients undergoing hypospadias surgery. Both analgesic techniques are safe. |
| Kendigele n et al 2016 | Turkey | November 2014 to May 2015 | 80 | I-II | Nerve stimulator-guided pudendal block | Bupivacaine 0.5 ml/kg 0.25% | Caudal block | 1 ml/kg dose of 0.2% bupivacaine | The Children Hospital of Eastern Ontario Pain Scale (CHEOPS) | Postoperative pain, Time to perform the block, and Intraoperative and postoperative analgesic requirement | It is proposed that the PNB, a widely accepted method frequently employed in adult patients, can offer effective, enduring, and optimal pain relief during the perioperative period in children undergoing hypospadias surgeries. Furthermore, PNB could be deemed suitable for outpatient procedures because of its low incidence of adverse effects. |
| Naja el al 2013 | Lebanon | July 2008 to October 2011 | 80 | I | Nerve stimulator-guided pudendal block | 0.3 ml/kg 0.25% bupivacaine | Caudal block | One milliliter per kilogram 0.25% bupivacaine | Objective pain/discomfort scale" (OPS) | Pain assessment, Time to perform the block, and Analgesics dosage. | Patients who received PNB had reduced analgesic consumption and pain within the first 24 hours postoperatively compared with CB. |
| Kundra et al 2012 | India | July 2008 to July 2010 | 53 | I-II | Penile block | Bupivacaine 0.25%, 0.5 mg/kg | Caudal block | 0.25% bupivacaine 0.5 ml/kg | VAS score | Duration of analgesia, Total oral paracetamol consumption, complication rate, and Pain assessment | Penile block provided better analgesia compared to caudal epidural block / postoperative urethral fistula formation was more likely in caudal block |

**Table 2.** Baseline characteristics of the participants. SD: standard deviation

| ID |  | Patient characteristics |  |  |  |  |  |  |  |  |  |  |
| --- | --- | --- | --- | --- | --- | --- | --- | --- | --- | --- | --- | --- |
|  |  | Population in each group |  | Patient's age, years |  |  | Weight,kgs |  |  | Height,cm |  |  |
|  |  | Sample Size | Total | Mean | SD | Total | Mean | SD | Total | Mean | SD | Total |
| Ahmed et al 2024 | Intervention | 34 | 68 | 7.06 | 2.4 | 34 | 22.35 | 5.2 | 34 | NA | NA | NA |
|  | Control | 34 |  | 6.78 | 2.12 | 34 | 20.6 | 4.5 | 34 | NA | NA | NA |
| Ahmed et al 2023 | Intervention | 30 | 60 | 3.7 | 1.96 | 30 | 15 | 5.8 | 30 | NA | NA | NA |
|  | Control | 30 |  | 3.5 | 1.75 | 30 | 14.8 | 5.06 | 30 | NA | NA | NA |
| Elbadry et al 2023 | Intervention | 41 | 81 | 2.95 | 1.07 | 41 | 15.9 | 2.11 | 41 | NA | NA | NA |
|  | Control | 40 |  | 2.75 | 1.35 | 40 | 14.93 | 2.37 | 40 | NA | NA | NA |
| Khalil et al 2022 | Intervention | 50 | 100 | 7.4 | 1.4 | 50 | NA | NA | NA | NA | NA | NA |
|  | Control | 50 |  | 7.3 | 1.6 | 50 | NA | NA | NA | NA | NA | NA |
| Choudhry et al 2022 | Intervention | 30 | 57 | 0.85 | 0.47 | 30 | 8.9 | 0.32 | 30 | NA | NA | NA |
|  | Control | 27 |  | 0.75 | 0.24 | 27 | 9.2 | 0.26 | 27 | NA | NA | NA |
| Karami et al 2021 | Intervention | 25 | 50 | 2.03 | 1.5 | 25 | 12.76 | 4.2 | 25 | 81.92 | 12.99 | 25 |
|  | Control | 25 |  | 2.3 | 1.45 | 25 | 13.28 | 3.73 | 25 | 82.4 | 11.99 | 25 |
| Ahmed et al 2021 | Intervention | 25 | 50 | 5.07 | 1.88 | 25 | 18.12 | 3.79 | 25 | 105.2 | 9.51 | 25 |
|  | Control | 25 |  | 5.15 | 1.89 | 25 | 18.32 | 3.87 | 25 | 105.24 | 9.52 | 25 |
| Kendigelen et al 2016 | Intervention | 40 | 80 | 4.38 | 2.77 | 40 | 19.15 | 9.22 | 40 | NA | NA | NA |
|  | Control | 40 |  | 3.79 | 2.33 | 40 | 16.79 | 6.34 | 40 | NA | NA | NA |
| Naja el al 2013 | Intervention | 40 | 80 | 3.1 | 1.1 | 40 | 15.3 | 2.8 | 40 | 92.6 | 14.3 | 40 |
|  | Control | 40 |  | 3.2 | 1.1 | 40 | 15.3 | 2.2 | 40 | 96.3 | 10.9 | 40 |
| kundra et al 2012 | Intervention | 27 | 53 | 6 | 2 | 27 | 18.9 | 5.5 | 27 | NA | NA | NA |
|  | Control | 26 |  | 7 | 2 | 26 | 19 | 5.9 | 26 | NA | NA | NA |

### 3.3. Risk of Bias

We used Cochrane RoB 2 to evaluate the risk of bias. Six studies had some concerns in the overall risk of bias [14,15,17,20,22,23], while no studies showed an overall high risk of bias. RoB results are shown in **(Figure 2-a).** In addition, the RoB decisions for each domain are outlined in **(Figure 2-b and Table S2).**

**Figure 2:**
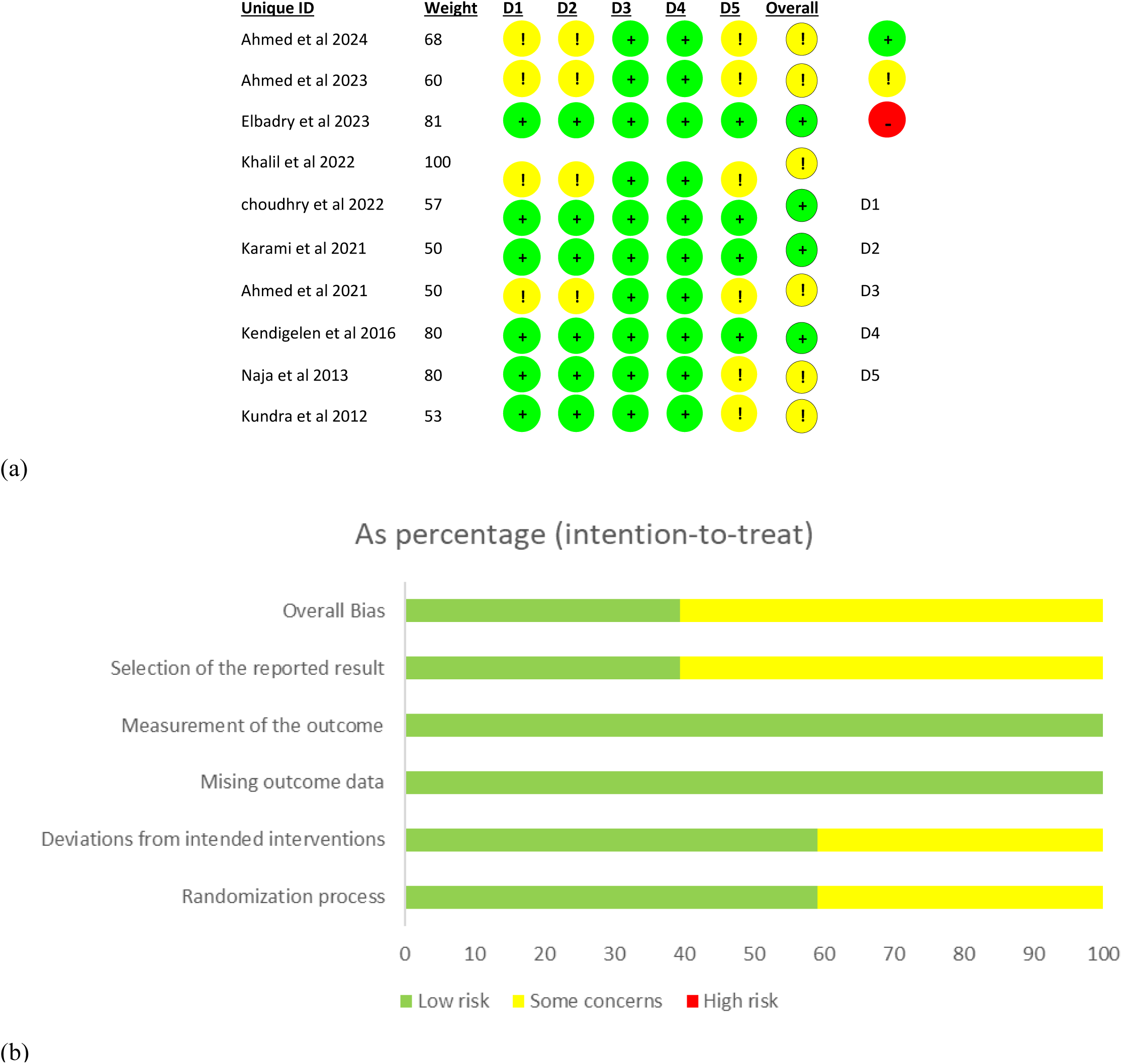
Quality assessment of the risk of bias, (a) Risk-of-bias tool 2 (RoB2) for each included study. (b) Overall risk of bias.

### 3.4. Primary Outcome

#### 3.4.1. Postoperative Pain Scores

##### 3.4.1.1. PNB vs. CB

PNB was significantly associated with lower pain scores compared to CB in 6, 12, and 24 hours using the random effect model (SMD: -0.50 with 95% CI [-0.77, -0.23], P=0.0003) **(Figure 3-a)**, (SMD: -1.95 with 95% CI [-3.29, -0.61], P=0.004) **(Figure 3-b)**, (SMD: -1.20 with 95% CI [-2.29, -0.11], P=0.03) **(Figure 3-c)** respectively. Pooled studies were significantly heterogenous (P = 0.003; I² = 75%), (P < 0.00001; I² = 96%), (P < 0.00001; I² = 94%) respectively. Heterogeneity was best resolved only in Pain 6 hours by excluding Kendigelen et al 2016 (P = 0.32; I² = 15%).

**Figure 3:**
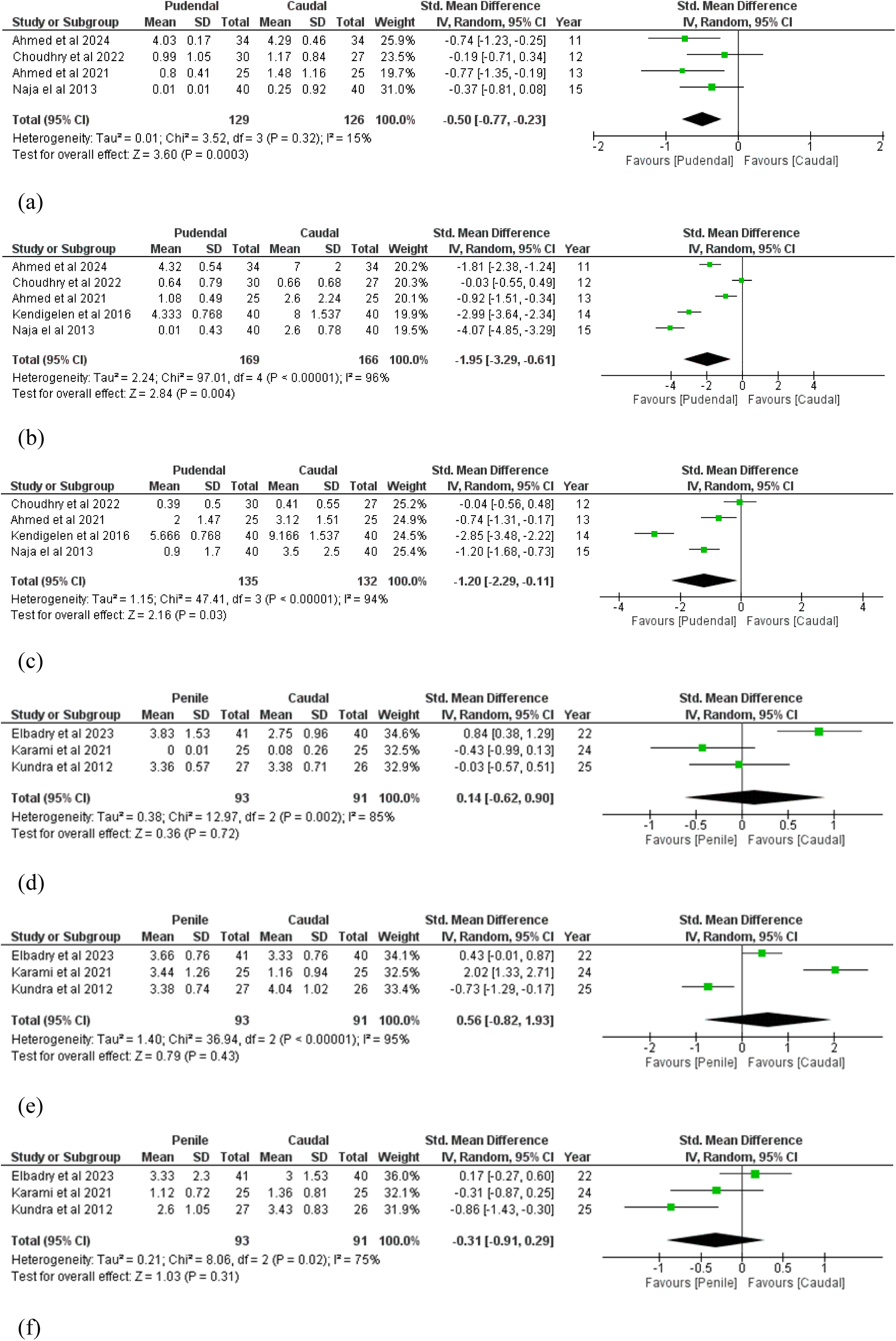
Summary of postoperative pain scores in both comparisons. Pudendal nerve block (PNB) vs Caudal epidural block (CB) in (a) 6 hours, (b) 12 hours, (c) 24 hours. Dorsal penile nerve block (DPNB) vs CB in (d) 6 hours, (e) 12 hours, (f) 24 hours. IV, Inverse variance; CI, Confidence interval; df, degree of freedom.

##### 3.4.1.2. DPNB vs. CB

DPNB did not show any significant difference compared to CB in terms of pain scores throughout all selected timepoints 6, 12, and 24 hours using the random effect model (SMD: 0.14 with 95% CI [-0.62, 0.90], P=0.72) **(Figure 3-d)**, (SMD: 0.56 with 95% CI [-0.82, 1.93], P=0.43) **(Figure 3-e)**,, (SMD: -0.31 with 95% CI [-0.91, 0.29], P=0.31) **(Figure 3-f)**, respectively. Pooled studies were significantly heterogenous (P = 0.002; I² = 85%), (P < 0.00001; I² = 95%), (P = 0.02; I² = 75%) respectively. Heterogeneity was not resolved by leave-one-out sensitivity analysis.

### 3.5. Secondary Outcomes

#### 3.5.1. PNB vs. CB

##### 3.5.1.1. Analgesic Consumption within 24 hours postoperatively

PNB was significantly associated with less analgesic consumption compared to CB (RR: 0.23 with 95% CI [0.05, 0.99], P= 0.05) **(Figure 4-a)**. Pooled studies demonstrated heterogeneity in analgesic consumption, which leave-one-out analysis did not resolve. (P < 0.00001; I² = 93%).

**Figure 4:**
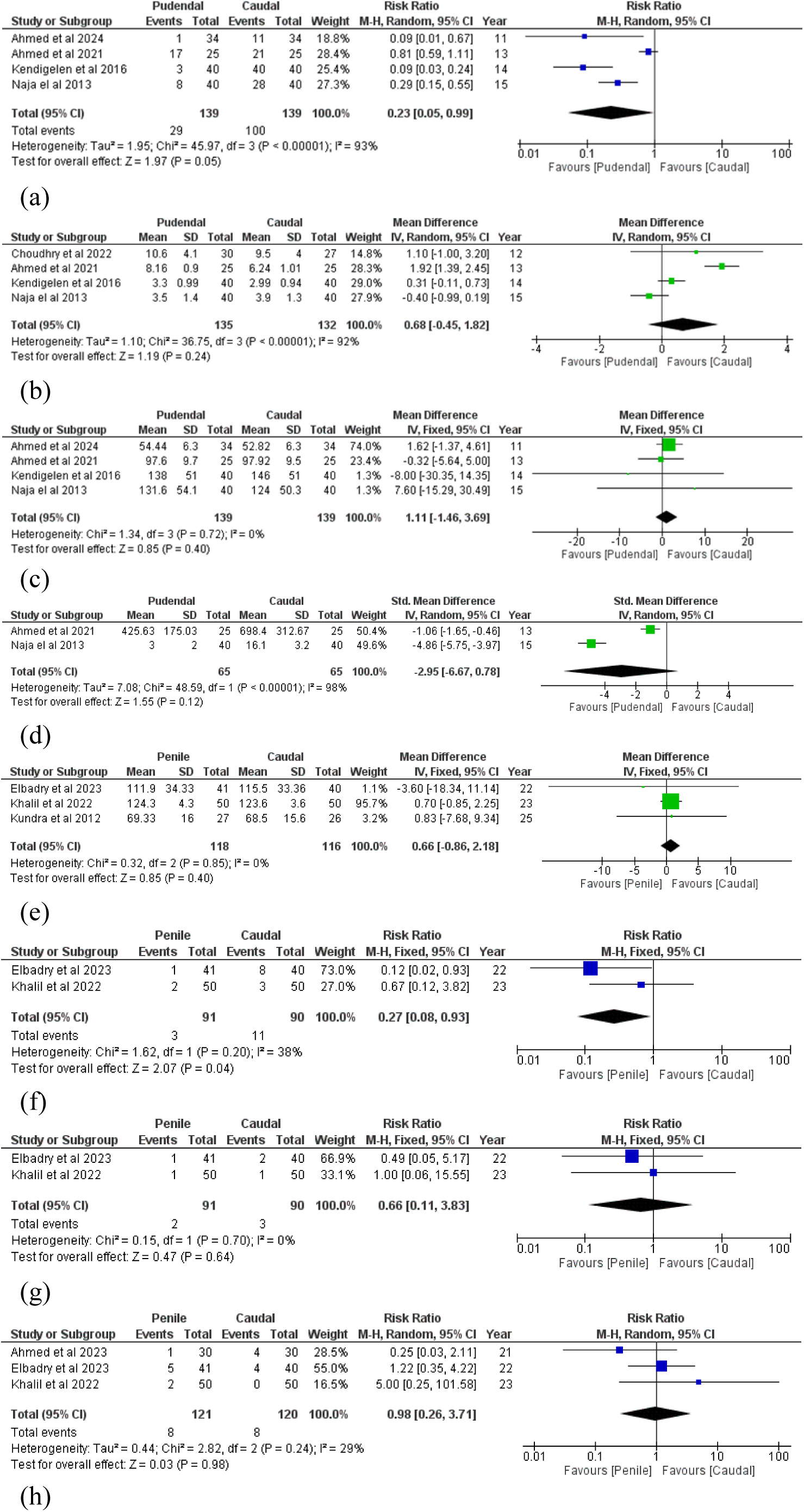
Summary of secondary outcomes in both comparisons. (a) Analgesic consumption in PNB vs CB. (b) Block procedure time in PNB vs CB. (c) Surgery time in PNB vs CB. (d) Total dose of acetaminophen in PNB vs CB. (e) Surgery time in in DPNB vs CB. Incidence of (f) hypotension, (g) bradycardia, (h) vomiting in DPNB vs CB. M-H, Mantel-Haenszel; CI, Confidence interval; df, degree of freedom; IV, Inverse variance.

##### 3.5.1.2. Block Procedure Time

Results showed no significant difference between the two studied groups in Block procedure time (MD: 0.68 with 95% CI [-0.45, 1.82], P=0.24) **(Figure 4-b)**. Pooled studies demonstrated heterogeneity in block procedure time, which leave-one-out analysis did not resolve. (P < 0.00001; I² = 92%).

##### 3.5.1.3. Surgery Time

There was no significant difference between PNB and CB in surgery time using the fixed effect model (MD: 1.11 with 95% CI [-1.46, 3.69], P= 0.40) **(Figure 4-c)**. The pooled studies were homogenous (P = 0.72; I² = 0%).

##### 3.5.1.4. Total Dose of Acetaminophen

Results showed no significant difference between the two studied groups in Block procedure time (SMD: -2.95 with 95% CI [-6.67, 0.78], P=0.12) **(Figure 4-d)**. Pooled studies demonstrated heterogeneity in total dose of acetaminophen, which leave-one-out analysis did not resolve. (P < 0.00001; I² = 98%).

#### 3.5.2. DPNB vs. CB

##### 3.5.2.1. Surgery Time

There was no significant difference between DPNB and CB in surgery time using the fixed effect model (MD: 0.66 with 95% CI [-0.86, 2.18], P= 0.40) **(Figure 4-e)**. The pooled studies were homogenous (P = 0.85; I² = 0%).

##### 3.5.2.2. Safety Outcomes

DPNB was significantly associated with a decreased incidence of hypotension (RR: 0.27 with 95% CI [0.08, 0.93], P= 0.04) **(Figure 4-f)**. However, there was no significant difference between DPNB and CB in the incidence of bradycardia (RR: 0.66 with 95% CI [0.11, 3.83], P= 0.64) **(Figure 4-g)**, and vomiting (RR: 0.98 with 95% CI [0.26, 3.71], P= 0.98) **(Figure 4-h)**. The pooled studies were homogenous in the incidence of hypotension (P = 0.20; I² = 38%), bradycardia (P = 0.70; I² = 0%), and vomiting (P = 0.24; I² = 29%).

## 4. Discussion

The aim of the present updated systematic review and meta-analyses was to assess the efficacy and safety of pudendal nerve block (PNB), dorsal penile nerve block (DPNB), and caudal block (CB) in postoperative analgesia as well as the rate of complications in pediatric hypospadias repair. The outcomes of our recent study demonstrate significant differences between these local anesthesia techniques, especially with respect to postoperative pain control as well as individual complications, with considerable heterogeneity found in different analyses.

Regarding postoperative analgesia, our meta-analysis indicates that PNB was associated with significantly lower pain scores compared to CB at 6, 12, and 24 hours postoperatively, and also resulted in significantly lower analgesic consumption within the first 24 hours. This suggests a potential superiority of PNB over CB for providing extended pain relief after hypospadias surgery.

The superior analgesic performance of PNB over CB in our study concurs with other studies, in that PNB allows for an extended sensory block of the penis. The pudendal nerve supplies the entire penis, including both its ventral skin and the frenulum through its perineal branch, whilst the cauda equina innervates a broader area of the saddle region. Therefore, blocking the pudendal nerve will provide more precise analgesic effects than caudal block due to its broader innervation distribution [5,24].

The results have partial concordance with the systematic review of Zhu et al. [25], who found that peripheral blocks (PNB and DPNB, combined in their pooled analysis of randomized controlled trials [RCT]) were associated with decreased pain scores at 24 hours when compared with CB. However, Zhu et al. failed to detect a statistically significant difference in the use of supplemental analgesia within the same time frame based on their limited RCT database. Moreover, Zhu et al. found that CB produced a significantly reduced duration of analgesia when compared with peripheral blocks in trials in RCT, which is in some contrast with our finding of reduced analgesic use in the PNB group. This would suggest that PNB is not just associated with decreased pain severity but may have a longer analgesic duration of action when compared with CB. Our finding of reduced need for rescue analgesia when PNB is used compared with CB adds credence to its desirability as an outstanding analgesic strategy in this surgery. Our analysis also showed great heterogeneity of pain results, suggesting that between-study differences in blocking method, anesthetic used, pain assessment strategy, or patient demographic might influence the outcome.

Compared with PNB, our analysis did not find statistically significant postoperative pain score differences between CB and DPNB at 6, 12, or 24 hours. This finding indicates that CB and DPNB may have similar analgesic efficacy in the first postoperative period after hypospadias repair. This finding is in line with the overall inferences from the randomized trials of Zhu et al. [25], who found that peripheral nerve blocks (including DPNB) did not show statistically significant variations in demand for rescue analgesia when compared with CB, in spite of the fact that these interventions had lower pain scores recorded for them at the 24-hour time interval. The large heterogeneity found in both our analysis as well as that of Zhu et al. with regard to pain outcomes attests to the heterogeneity of outcomes between studies, possibly due to variations in study design, pain-measurement methods, dosages of administered anesthetics, or different demographic profiles of patients.

The safety profiles of these regional anesthetic methods, particularly the controversial relationship of CB with postoperative surgical complications like urethrocutaneous fistula (UF) and glans dehiscence (GD), warrant meticulous analysis. Our study focused on other safety measurements, rather than directly measuring the incidence of UF or GD. Our results showed that the use of DPNB was associated with a significantly decreased incidence of hypotension when compared with CB, while no statistically significant differences were found concerning bradycardia or vomiting between the two blocks. This finding indicates a potential hemodynamic advantage of DPNB over CB.

However, the broader literature presents conflicting evidence. Early meta-analyses by Tanseco et al. [26] and Goel et al. [27] reported a significantly higher risk of UF and overall complications associated with CB compared to NCB (including PB or no block). Tanseco et al. [26], in their meta-analysis primarily of observational studies, found an overall increased risk of complications with CB, particularly for proximal hypospadias. Goel et al. [27] found an RR of 1.81 for UCF with CB. These findings fueled concerns that CB-induced vasodilation and penile engorgement might impair wound healing.

Conversely, more recent large-scale meta-analyses by Adler et al. [28] (10 studies, 3201 patients) and Xia et al. [29] (16 studies, 3855 patients) found no significant association between CB and the incidence of UCF or GD compared to NCB (PB or GA). Adler et al. reported an RR of 1.11 (95% CI 0.88-1.41) [28], and Xia et al. found an OR of 1.40 (95% CI 0.88–2.23) [29]. Subgroup analyses in Xia et al.’s study, stratified by study type, meatal location (distal only), type of NCB (PB or GA), surgeon/technique, and anesthetic dose, consistently showed no significant association [29]. These later meta-analyses included larger, more recent studies, potentially providing a more robust estimate of the effect. Adler et al.’s own large cohort study (n=983) also found no association, identifying proximal hypospadias severity, surgical duration, and age as significant predictors of complications, but not block type. Furthermore, the proposed mechanism of CB-induced penile engorgement has been questioned, with an ultrasound study showing no significant change in penile blood flow after CB [30].

Our results add depth to the complex understanding of the subject. While notable variations in operation duration between the block types were not encountered, the decreased risk of associated hypotension in DPNB compared with CB is worthy of specific note. CB has been thought to cause sympathetic blockage with consequent hemodynamic change; such impacts are typically weaker in the younger pediatric age group [30]. The lowered risk associated with DPNB, as a relatively more localized technique, could hold advantages for individual populations of patients. The clinical importance of such an observation, though, warrants further investigation, as post-operative hypotension was rarely documented as an immediate major outcome in various hypospadias studies prioritizing complications of the operation.

The variation in research results regarding CB and complications of surgery highlights the great challenges faced when attempting to interpret the current body of evidence, much of which is based on observational studies open to bias and confounding variables. As explained by Braga et al. [31] and Adler et al. [30], the variables of severity of hypospadias, the method of surgery used, as well as the skill of the surgeon, are important confounders that may influence rates of complications irrespective of the anesthetic technique used. The selection bias phenomenon in which patients with more severe hypospadias will be treated with CB could explain the correlation found in some studies [26,31]. The outcomes demonstrated by Xia et al. [29], Adler et al. [28], Goel et al. [27], as well as observational data analysis by Zhu et al. [25], reveal no correlation in terms of significance after an effort is made to control for individual variables, thereby supporting the impression that CB is not the major influence on rising UF or GD rates. Our observation of reduced hypotension with DPNB compared with CB adds another factor to the safety analysis, with the benefits of DPNB in this specific regard.

One of the key limitations found in our results was the extensive heterogeneity found in the meta-analyses on primary outcomes, i.e., pain scores, as well as secondary outcomes, i.e., analgesic use. The same heterogeneity was also the key feature in the meta-analyses of Zhu et al. [25], Tanseco et al. [26], Goel et al. [27], and Adler et al. [28]. The variability is presumably because of variations in research methods, anesthetic techniques (type of drug, drug concentrations, drug amount), surgical techniques, definitions of outcome, as well as types of patients in studies included. The low-to-moderate quality of the evidence, including from observational studies as Tanseco et al. [26] brought out, as well as the bias as presented in the discussions of Braga et al. [31] and Adler et al. [30], adds complexity in the integration of results as well as in drawing ultimate conclusions.

Therefore, the results synthesized in our review, when compared with similar studies, suggest that PNB can confer superior postoperative analgesia compared with CB for hypospadias repair in the pediatric age group. DPNB also has analgesic profiles comparable with CB, but possibly with an advantage in terms of safety with respect to postoperative hypotension. The association of CB with postoperative complications, such as UF and gastric distension GD, remains controversial; while some analyses suggest an association, in particular in observational studies of more complex types of hypospadias [26], other meta-analyses that control for confounders demonstrate no such association [25, 27-2]. As such, the choice of the anesthetic block must consider the desired trade-off between analgesic efficacy and potential side effects, while recognizing the profound variability and methodological limitations of the literature [30-31]. There is an urgent need for large, randomized, well-delineated trials comparing directly the use of PNB, DPNB, and CB with careful control of both hypospadias severity and techniques used, in order to finally determine the relative benefits of these techniques.

## 5. Conclusions

Our study showed CB to be more associated with complications compared to DPNB, with less postoperative analgesia compared to PNB; thus in hypospadias repair surgery, there should be more consideration of regional block methods of PNB, which showed more efficacy, and DPNB, which provided more safety. However, regional block selection should balance analgesic efficacy with safety profile. More clinical trials that provide homogenous and no biased data is needed.

## Data Availability

All data produced in the present work are contained in the manuscript

## 6. Statements and declarations

## 6.1. Acknowledgements.

The authors would like to thank Mohamed Abd-ElGawad and Mahmoud Noureddine Srouji for their support regarding this article.

## 6.2. Authors Contributions

M.A.: Conceptualization, Data curation, Investigation, Methodology, Project administration, Resources, Validation, Writing – review and editing. O.C.A.: Data curation, Investigation, Methodology, Writing – review and editing. M.J.A.: Formal analysis, Software, Visualization, Writing – original draft, Writing – review and editing. A.A.A.: Data curation, Investigation, Writing – original draft. M.A.K.: Data curation, Investigation, Writing – original draft. A.M.: Data curation, Investigation, Writing – review and editing. K.A.: Data curation, Investigation, Writing – review and editing. M.A.A.: Writing – original draft, Writing – review and editing. M.T.S.: Supervision, Writing – review and editing. M.M.: Conceptualization, Writing – review and editing. All authors read and approved the final manuscript.

## 6.3. Funding

This research did not receive any specific grant from funding agencies in the public, commercial, or not-for-profit sectors.

## 6.4. Declarations of interest

None.

## 6.5. Ethical approval and patient consent

Not applicable.

## 6.6. Availability of data and materials

Not applicable.

## Notes

### Competing Interest Statement

The authors have declared no competing interest.

### Funding Statement

This study did not receive any funding

