## Supplemental tables for "Analgesic Efficacy and Safety of Caudal Epidural, Dorsal Penile Nerve, and Pudendal Nerve Blocks in Hypospadias Repair Surgery in Children: Updated Meta-Analyses"

**Title.**

**Author names and affiliations.**

Moumen Arnoaut^a*^, Omar Chikh Amine^a^, Mohamad Jaber Arab^a^, Abdullah Aladnan Aljammas^a^, Mohamad Amin Kreid^a^, Abdulmajid Makki^a^, Kais Alissa^a^, Mohammed Al-mahdi Al-kurdi^a^, Mohammad Tammam Saffour^b^, Mohamad Morjan^c^.

^a^ Faculty of Medicine, Aleppo University, Aleppo, Syria.

^b^ Department of Anaesthesia and Intensive Care Medicine and Pain Management, Faculty of Medicine, Aleppo University, Aleppo, Syria.

^c^ Department of Pediatric Surgery, Faculty of Medicine, Aleppo University, Aleppo, Syria.

**Corresponding author.**

Moumen Arnaout

Faculty of Medicine, Aleppo University, Aleppo, Syria.


(Pudendal OR Penile OR DPNB OR "Dorsal Nerve" OR Caudal OR Epidural) AND (Hypospadias OR Hypospadia)
____________________________________________________________________________________

| **Database** | **Search Terms** | **Search Field** | **Search Results** |
| --- | --- | --- | --- |
| Pubmed | (Pudendal OR Penile OR DPNB OR "Dorsal Nerve" OR Caudal OR Epidural) AND (Hypospadias OR Hypospadia) | All Field | 3908 |
| CENTRAL | (Pudendal OR Penile OR DPNB OR "Dorsal Nerve" OR Caudal OR Epidural) AND (Hypospadias OR Hypospadia) | All Field | 293 |
| WOS | **ALL=(**(Pudendal OR Penile OR DPNB OR "Dorsal Nerve" OR Caudal OR Epidural) AND (Hypospadias OR Hypospadia)**)** | All Field | 1905 |
| SCOPUS | **TITLE-ABS-KEY (** ( pudendal OR penile OR dpnb OR {dorsal nerve} OR caudal OR epidural ) AND ( hypospadias OR hypospadia ) **)** | Title, Abstract, Keywords | 2507 |
| EBSCOhost | (Pudendal OR Penile OR DPNB OR "Dorsal Nerve" OR Caudal OR Epidural) AND (Hypospadias OR Hypospadia) | All Field | 666 |
| Clinicaltrials.gov | (Pudendal OR Penile OR DPNB OR "Dorsal Nerve" OR Caudal OR Epidural) AND (Hypospadias OR Hypospadia) | All Field | 119 |
| International Clinical Trials Registry Platform (ICTRP) | (Pudendal OR Penile OR DPNB OR "Dorsal Nerve" OR Caudal OR Epidural) AND (Hypospadias OR Hypospadia) | All Field | 61 |

**Table S1**: Search strategy across databases and registries.

| **Study** | **Randomization process** | **Deviations from intended interventions** | **Missing outcome data** | **Measurement of the outcome** | **Selection of the reported result** | **Overall Bias** |
| --- | --- | --- | --- | --- | --- | --- |
| Ahmed et al 2024 | Some concerns | Some concerns | Low | Low | Some concerns | Some concerns |
| Ahmed et al 2023 | Some concerns | Some concerns | Low | Low | Some concerns | Some concerns |
| Elbadry et al 2023 | Low | Low | Low | Low | Low | Low |
| Khalil et al 2022 | Some concerns | Some concerns | Low | Low | Some concerns | Some concerns |
| choudhry et al 2022 | Low | Low | Low | Low | Low | Low |
| Karami et al 2021 | Low | Low | Low | Low | Low | Low |
| Ahmed et al 2021 | Some concerns | Some concerns | Low | Low | Some concerns | Some concerns |
| Kendigelen et al 2016 | Low | Low | Low | Low | Low | Low |
| Naja et al 2013 | Low | Low | Low | Low | Some concerns | Some concerns |
| Kundra et al 2012 | Low | Low | Low | Low | Some concerns | Some concerns |

**Table S2** Description of risk of bias (ROB2) assessment.
